# Oral Healthcare Providers’ Well-being, Health and Experience (WHE) in Dental Care Settings: A Protocol for an Umbrella Review of Systematic Reviews and Meta-analyses

**DOI:** 10.1101/2025.07.30.25332274

**Authors:** Pascaline Kengne Talla, Bile Yacouba Djedou, Geneviève Gore, Belinda Nicolau, Paula Bush

## Abstract

**Objectives:** Over the decades, healthcare systems have faced significant pressures in supporting the well-being, health and experiences (WHE) of oral healthcare providers (OHCPs), while ensuring high quality of care. Emerging evidence reports physical, mental and social challenges among OHCPs, shaped by multilevel factors; however, a comprehensive synthesis of these issues is still needed. This umbrella review aims to summarize evidence on the barriers and enablers to OHCPs’ WHE in dental care settings.

**Methods:** The study will follow the Joanna Briggs Institute and Cochrane Handbook guidelines for umbrella reviews. The populations, intervention, context, outcomes and study design (PICOS) are defined as the following: (P) oral healthcare professionals, (I) definitions of WHE from authors, (C) any country or setting, (O) enablers and barriers on OHCPs’ WHE, (S) systematic reviews with and without meta-analysis. We will conduct the search strategy through six databases, including MEDLINE, Embase, CINAHL, Web of Science, PsychINFO, Cochrane Library, along with grey literature. Two independent reviewers will screen and select relevant reviews and extract data from systematic reviews and meta-analyses. We will assess methodological quality using the AMSTAR-2 and ROBIS tools. We will use the GROOVE tool for overlapping. We will synthesize barriers and enablers using the Consolidated Framework for Implementation Research. We will present the results in diagrammatic and tabular, along with a narrative summary.

**Results and conclusion:** The initial MEDLINE search identified 492 studies. The study findings will consolidate evidence on determinants impacting OHCPs’ WHE, providing insights for key stakeholders to foster safer and healthier dental care settings.

**Key Points:** **What is already known on this topic**

Oral healthcare system is constantly changing and adapting to meet emerging social priorities and to deliver high quality and patient-centered care. As gatekeepers of the healthcare system, OHCPs’ WHE is essential to achieve the quintuple aim. Poor OHCP’s WHE is influenced by a range of factors, resulting in dental errors, adverse events, wastage in care and high costs.

**What this study adds**

This first umbrella review will summarize the multiple factors influencing positively and negatively OHCP’s WHE using the Consolidated Framework of Implementation Research, a robust and solid framework in implementation science. It will provide a better understanding of the challenges faced by OHCPs regarding their WHE and its impacts on a high quality of oral health care delivery.

**How did this study might affect research, practice and policy**

Understanding the importance of multiple levels (e.g., individual, contextual, and systemic) factors on OHCP’s WHE will provide valuable insights for decision makers, researchers, patients and guideline developers and OHCPs on OHCP’s WHE in developing successful implementation strategies to improve healthy work conditions to maximize their joy, and ultimately patient oral healthcare outcomes and experiences.

## Introduction

Over the decades, oral healthcare systems have faced significant challenges and pressures to retain their healthcare providers, while addressing issues of shortage, turnover, burnout, fatigue, addiction and suicide.^1-4^ These challenges have caused severe oral health inequalities by disproportionally affecting the marginalized people, as those living in remote and rural regions, people with disabilities, indigenous communities and aging population.5 The oral healthcare systems must adapt to meet the emerging societal priorities as the demographic shifts, with the comorbidities and complex chronic diseases and conditions, along with the development of technological innovations and the environmental sustainability issues in dentistry. However, strengthening the oral healthcare system to deliver high quality, safe, sustainable and patient-centered care depends on oral healthcare providers (OHCPs)’ well-being, health and experiences (WHE) according to the Quintuple aim6 and the value-based oral healthcare movement.^7^ Poor OHCPs’ WHE contributes to increased risk of dental errors, adverse patient events and increased avoidable costs.^8, 9^ As gatekeepers of the healthcare system,^10^ OHCPs’ WHE is essential to enhance patient outcomes and experiences, improve the population oral health and reducing costs of oral healthcare.^11, 12^

Several primary studies and systematic reviews have addressed major components of WHE among OHCPs, including burnout,^4^ job satisfaction,^13^ stress,^14^ depression,^15^ professional career,^8^ weight stigma^16^ and physical and mental health.^17, 18^ The increased burnout has been exacerbated during the COVID-19 pandemic, with 71% of OHCPs affected.^19^ The fear of COVID-19 infection, increased workload, practice put on hold, loss of income resulting from further lockdowns, the cost of personal protective equipment, and patient volume decline were among the most contributing factors. Inequalities about burnout levels are higher among female dentists, young dentists (18-34 years versus 55 years and over), dental specialists, those working in residential areas and those in public health centers compared men (66 % versus 50%), dental specialists practicing in private office or academic settings, and general dental practitioners.^19, 20^ Dental education’s coursework, student debts, and undervaluation further demotivate students and professionals.^20^ Furthermore, OHCPs are among high-risk occupational group for hearing loss due to high and continued exposure to noise and musculoskeletal disorders due to posture in their workplace.^21^ The prevalence of these disorders are variable (between 60% and 100%)^18^ with common issues like back problems, arthritis, and job-related discomfort, particularly in the back, and shoulders.^21^

To tackle the ongoing crisis in dentistry, it is essential to deepen the understanding of the diverse stressors related to business pressures, clinical challenges, healthcare regulations, work environment and conditions, and societal expectations, often outside OHCPs’ locus of control, that significantly influence their WHE.^2^ For instance, the factors that impede or enable OHCPs’ WHE may be located at macro (e.g., public policy, institutional support regulatory structures), meso (e.g., professional dynamic, interactions within the dental team, colleagues, patients and families, dentistry as a profession, and integration of new technologies) and micro (e.g., socio-cultural factors, income, personal fulfillment, professional career) levels.^4, 8, 13, 15, 22^ In a rapid review on key determinants of health and well-being of dentists within the United-Kingdom, authors reported that workplace characteristics were most often explored in the literature, along with a gap about the underlying strategies to improve the health and well-being of dentists.^3^ Drawing on knowledge mobilization, implementation science, and behavioral change principles, leveraging theory becomes pivotal in guiding the design and the implementation of tailored interventions and strategies to improve the delivery of high quality oral healthcare.^23^ These implementation efforts will be useful to promote the successful and sustainable adoption of innovations while addressing what matters most to the OHCPs’ WHE and the needs of their patients/families. However, it is crucial to better understand the underlying determinants of OHCPs’ WHE as they will improve safe, efficient and equitable oral healthcare.

While previous systematic reviews (SRs),^8, 14, 17, 22^ and overviews of SRs and systematic reviews and meta-analyses (SRs, and SR-MAs).^18, 24^ have examined certain aspects of OHCPs’ WHE and its determinants, a comprehensive synthesis of evidence on these determinants is needed due to the complexity of this concept. To this end, this umbrella review will provide a synthesis of the existing literature on factors that influence positively or negatively OHCPs’ WHE in delivering oral healthcare. A preliminary search in MEDLINE, the Cochrane Database of Systematic Reviews, and Joanna Briggs Institute Evidence Synthesis revealed no published or in-progress overviews, umbrella reviews or systematic reviews of systematic reviews and meta-analysis on determinants of OHCPs’ WHE. No similar review protocol is registered in PROSPERO or Open Science Framework. The findings of this umbrella review will provide insights to inform decision-makers, OHCPs, patients, and guideline developers on the factors that contribute to and impact OHCPs’ WHE, as well as to identify knowledge gaps to inform future research and directions.

### Purpose

Our objective is to summarize evidence from systematic reviews and meta-analyses (SRs, and SR-MAs) on the barriers and enablers to OHCPs’ WHE in dental care settings.

### Research questions

To achieve this objective, the following questions will be addressed:

1. What multilevel (e.g., individual, contextual and systemic) factors impact OHCPs’ WHE and how do these factors exert their effects?

2. What are the components of OHCPs’ WHE (e.g., job satisfaction, work engagement, burnout) and the most common determinants?

3. What are the implementation theories, models, and frameworks that have been used to guide research in these areas?

## Methods

This protocol was registered in the International Prospective Register of Systematic Reviews (PROSPERO CRD42024591840). The review will be conducted according to the Joanna Briggs Institute Manual for Evidence Synthesis for umbrella reviews,^25^ and the Cochrane Handbook for an overview of reviews.^26^ We will report it in accordance with the Preferred Reporting Items for Systematic Reviews and Meta-Analyses Protocols (PRISMA-P) statement.^27^ This study began on July 13^th^, 2024 and will end on December 30^th^, 2025. However, all modifications to the protocol will be recorded and described in the final umbrella review report. The PRISMA checklist is available as Appendix 1.

### Eligibility criteria

We will use the “PICOS” format: Participants, Intervention or concept, Context, Outcome, and Study Design.

### Participants/population

We will include systematic reviews, systematic reviews with meta-analyses reporting of any oral healthcare providers (e.g., dentists, dental specialists, dental hygienists, dental assistants, dental prothesis experts, dental nurses, dental auxiliaries, dental therapists, undergraduate and post-graduate dental students and dental hygienists, etc.).

### Concept/Intervention

To be considered in the umbrella review, all SRs must provide a definition of the concepts of WHE. While clinician’s WHE is broad concept, with the lack of agreement and the inconsistencies on its definitions,^28^ there are some attempts to define this complex concept. Drawing on previous definitions, ^29, 30^ we will refer to OHCPs’ WHE as positive experiences across all job-related aspects, enabling OHCPs to reach their full potential within dental care settings and improve their physical, emotional and social health and quality of life. There include some features without exhaustivity: i) emotional factors such as stress levels, burnout, anxiety, depression, resilience, fatigue; ii) job satisfaction/performance/engagement/security, professional fulfillment, workload, and musculoskeletal problems; iii) interactions with patients/families and colleagues and balance between personal and professional life. SRs reporting on the efficacy or effectiveness of interventions (e.g., educational, training, regulatory, financial) aiming to improve OHCPs’ WHE will be excluded. In addition, SRs that emphasize relevant features such washing hands, allergies, gloves will be excluded, despite their potential impacts on OHCPs’ WHE.

### Context

We will consider SRs that include participants from all countries, regardless of the country’s income level) and from all geographical characteristics (e.g., urban, rural, and remote regions). Additionally, all dental settings will be considered, including private and public clinics, dental schools, hospitals, mobile dental clinics, community health centers, and long-term care facilities.

### Outcomes

The outcomes of these studies are barriers and enablers to OHCPs’ WHE.

### Types of studies

We will include SRs and SR-MAs. They could include any type of primary studies. A SR is defined as a reproducible, standardized, and transparent approach aimed at identifying, evaluating, and summarizing evidence from primary studies on a specific topic.^31^ To be considered in this umbrella review, all SRs must have conducted searches in at least two databases and performed quality assessments.^31^

### Information sources, search strategy, and data management

A comprehensive search strategy was developed interactively by an expert librarian (GC) with the support of the research team. The search strategy combines controlled vocabulary terms (e.g., MeSH, Emtree) and keywords to represent terms related to oral health care, work-related factors, job satisfaction, burnout, mental health, and patient-clinician interactions. It was initially developed in MEDLINE (Ovid) (Appendix 2) and will be adapted and run in the following additional databases: Embase (Ovid), CINAHL (EBSCO), Web of Science Core Collection (All Indexes), PsychINFO (Ovid), Cochrane Reviews. The (Cochrane Library/Wiley, Epistemonikos, Sociological Abstracts (ProQuest), Academic Search Complete (EBSCO), and ProQuest Dissertations and Theses Global (ProQuest). We will also perform searches in Sociological Abstract (ProQuest), Academic Search Premier (EBSCO), and ProQuest Dissertations & Theses, as well as in grey literature, including reports from governments and non-governmental organizations and databases such as Open Grey and the Agency for Healthcare Research and Quality. The initial MEDLINE search was conducted from its inception to November 2024, identifying a total of 492 SRs (see Appendix 2). No restrictions on publication dates or languages will apply. Reviews written in languages other than French or English will be translated using DeepL.^32^ The search strategy will be updated during the synthesis phase to identify any new relevant reviews published subsequent to the initial search.

### Study selection process

Identified records will be imported into Covidence, a systematic review management software. The research team will discuss the selection criteria and develop a selection tool. Then, a calibration exercise will be conducted (PKT and BYD) on 10% of the identified SRs to ensure a common understanding of the inclusion and exclusion criteria. The data selection process will follow three steps: 1) the reviewers will conduct a pilot selection of studies on 10% of the total of retrieved records. This pilot aims to facilitate a shared understanding of eligibility criteria by reviewers, and to clarify these criteria and refine them if needed; 2) after a conclusive pilot step, reviewers will independently screen studies based on titles and abstracts, rating them as either “include”, “exclude” or “unclear”; 3) reviewers will then independently screen full texts of all publications rated as “include” or “unclear”. This step of selection will also be rated “include”, “exclude”, or “maybe”. Disagreements at each stage will be resolved through discussion or consultation with a third reviewer. We will adjust the grid following this calibration exercise. All studies rated “include” or “unclear” will be considered for the second phase (i.e., full-text review). Decisions and reasons for exclusion will be recorded in Covidence.

### Data extraction

The data from each included SR will be extracted by two independent reviewers (PKT and BYD) using a predefined data extraction form, developed by the research team according to the JBI website resources for umbrella reviews^25^. Reviewers will perform a conclusive data extraction pilot on some studies before the main data extraction. This form will be designed to ensure that all relevant information to the objectives of this review is systematically collected. Disagreements between reviewers during data extraction will be resolved by consensus, or by consultation with a third author (PB). In case of missing data or the need for additional information, the authors of the relevant reviews will be contacted. If possible, the reviewers will not extract data from studies in which they have participated to avoid any conflict of interest and maintain the objectivity of the analysis. The following information will be extracted: i) review characteristics (e.g., first author, year of publication, language, country of the first author, published protocol, number of databases, type of review with or without meta-analysis, dental care settings, funding), ii) participants: characteristics and number; iii) components or dimensions of WHE; iv) barriers and enablers; v) methodological characteristics (e.g., use of theories, models or frameworks, quality assessment (e.g., tool used, results); additional data analysis approaches such as subgroup analyses, sensitivity analyses; certainty of evidence and heterogeneity). Heterogeneity will be quantified with Higgins’ I^2^; vi) other details: main conclusions, limitations, and conflicts of interest. For SR-MAs, effect sizes (e.g., rate ratio, risk ratio, odds ratio, mean difference or standardized mean difference for continuous data) as well as corresponding 95% CIs and *P* values will be extracted.

### Quality assessment

The quality assessment of SRs is a key component when conducting an overview of systematic reviews.^31^ It includes both evaluation of methodological quality and assessment of risk of bias, which are two distinct concepts.^33^ The methodological quality assessment focuses on ensuring adherence to the highest standards in the conduct and reporting of the research process,^33^ while the risk of bias assessment examines potential issues with the study design, execution, analysis, interpretation, or reporting that could affect the study’s results.^33^ To carry out this assessment, we will use two reliable and complementary tools: the AMSTAR-2 checklist (Assessment of Multiple Systematic Reviews 2) to evaluate methodological quality, ^33^ and the ROBIS tool to comprehensively assess the risk of bias.^33^ Despite the relevance of these tools, they are mostly for SRs of quantitative reviews and there is a lack of critical appraisal tools to assess the quality of SRs of qualitative studies.^34^ In fact, some criteria from AMSTAR-2 and ROBIS tools are not adapted for quality assessment of factors such as risk of bias, publication bias, heterogeneity, meta-analysis in SRs of qualitative research ^34^. However, these tools will be used within the limit of their scope.

The AMSTAR-2 tool, a comprehensive instrument designed to evaluate the key aspects of systematic reviews of randomized controlled trials (RCTs) and non-randomized trials (non-RCTs), consists of 16 items, including 7 critical domains (items 2, 4, 7, 9, 11, 13, and 15). Responses are categorized as: “yes” (item/question fully addressed), “no” (item/question not addressed), or “partial” (item/question partially addressed). The overall confidence in the review’s results is then classified into four levels: high, moderate, low, or critically low.^33^

The ROBIS tool is particularly effective at identifying sources of bias throughout the review process, such as study selection, data extraction, and result synthesis.^33^ It is structured around 24 questions organized into three phases: phase 1 (optional) assesses relevance; phase 2 identifies concerns with the review process; and phase 3 assesses the risk of bias in the review itself. Questions in phases 2 and 3 are answered using the following options: yes, probably yes, probably no, no, or no information. Concerns raised in these phases are categorized as low, high, or unclear. We will focus on phases 2 and 3 to establish an overall risk of bias for each review. Phase 2 examines concerns across four domains: study eligibility criteria, study selection, data collection and appraisal, and synthesis and conclusions. Each domain includes signaling questions and an assessment of bias risk, judged as low, high, or unclear. Phase 3 then summarizes the concerns identified in phase 2.^33^ Two independent reviewers will perform the quality assessment. Disagreements between the reviewers will be resolved through discussion or consultation with a third author.

### Overlap between studies

Overlap is a common concern in conducting SRs and overviews of SRs.^35^ The overlap refers to a duplication of eligibility criteria, which has not been reported as an update nor a replication.^35^ Following these recommendations, the overlap among included SRs will be visualized using the GROOVE (Graphical Representation of Overlap for OVErviews) tool,^36^ which provides a matrix of evidence indicating the number of primary studies and included systematic reviews. Additionally, it is important to estimate the degree of overlap using the Corrected Covered Area (CCA). A CCA of 0 to 5% indicates slight overlap, 6 to 10% moderate overlap, 11 to 15% high overlap, and above 15% very high overlap.^36^

### Assessment of the level of credibility

We will report the certainty of evidence assessments as presented in SRs by authors. For authors using GRADE (Grading of Recommendations Assessment, Development and Evaluation) approach for quantitative evidence,^37^ the quality of evidence will be classified as high, moderate, low, or very low according to four factors such as risk of bias, inconsistency, indirectness, imprecision, and publication bias. For authors using GRADE-CERQual^38^ to assess confidence in the evidence from qualitative evidence synthesis, we will report results across four domains: (a) methodological limitations, (b) coherence and (c) adequacy of data, and (d) relevance. The scoring system ranges from substantial concerns (one point) to no concerns to very minor concerns (4-points). These findings will be useful to identify which factors are supported by evidence with the highest level of confidence, and their corresponding level of evidence.

### Data synthesis

We will organize data extracted from the included SRs and SR-MAs into diagrams and tables. Additionally, we will conduct a narrative synthesis accompanying the tabular and diagrammatic presentations, summarizing the findings and discussing their implications. Quantitative results will be compared and integrated with qualitative results to address the umbrella review’s research questions and a depth-understanding of the implications of different contexts for OHCPs.

The barriers and enablers will be synthesized using the Consolidated Framework for Implementation Research (CFIR).^39^ CFIR is a meta-theoretical framework providing a repository of standardized implementation-related constructs at the individual, organizational, and external levels that can be applied across the spectrum of implementation research. CFIR contains 48 constructs and 19 subconstructs representing determinants of implementation across five domains: Innovation (OHCPs WHE), Outer Setting (e.g., national policy context), Inner Setting (e.g., dental clinic), Individuals (e.g., OHCP’s and patient’s motivation) and Implementation Process (e.g., assessing context).^39^ All SRs will be independently coded by two reviewers. Any disagreements that arise between the reviewers will be resolved through discussion, or with a third reviewer.

## Discussion

This umbrella review aims to consolidate evidence on OHCPs’ WHE in dental settings by including a synthesis of quantitative, qualitative, and mixed methods of systematic reviews. Our results will indicate which factors hinder and enable OHCPs’ WHE such as burnout, job satisfaction, work engagement, depression, musculoskeletal diseases and conditions. Furthermore, this umbrella review will provide valuable insights regarding how barriers and enablers to OHCPs’ WHE varies across different dental settings, geographical regions, the evidence supporting these findings and the most common implementation theories, models and frameworks used to study this topic. This holistic perspective on determinants factors is essential to an in-depth understanding of how context-specific variables may influence the generalizability of findings.

Strengths of this umbrella review include the use of a robust implementation framework (CFIR 2.0) to categorize multilevel factors influencing OHCPs’ WHE, the registration of the title, the assessment of methodological quality using two complementary tools, and the assessment of the overlap. To our knowledge, that is the first umbrella review on determinants of OHCPs’ WHE. Interestingly, mapping barriers and enablers with CFIR 2.0 is valuable for designing interventions and aligning them with relevant implementation strategy taxonomies, such as the Expert Recommendations for Implementing Change (ERIC).^40^ OHCPs’ WHE may affect their decision-making, their turnover, workforce sustainability, their performance, patient safety and the quality of care. This umbrella review aims to enhance our understanding of the complex factors involved in preventing, maintaining, increasing, or OHCPs’ WHE. It is also useful in identifying knowledge gaps to guide further research on strategies for ensuring sustainable healthcare systems. However, our umbrella review is associated with some limitations, including being limited to the inclusion of SRs conducted in at least two databases, and having conducted a quality assessment, while some papers qualifying as SRs, literature reviews can have relevant results.

To maximize the reach and impact of our results, our Knowledge Mobilization (KM) plan will include: i) publications in peer-reviewed academic journals such as the Journal of Occupational Health, and other journals in health services, dentistry, learning health system or implementation science; ii) presentations at local, national and international scientific events to raise awareness among a wide range of researchers, clinicians, patients and policy-makers about our results and recommendations; and iii) through social media platforms such as X, BlueSky, LinkedIn. The information generated by the umbrella review can be used to enhance both clinical practices, the delivery of high quality of oral healthcare and the OHCPs’ health and well-being, contributing to strengthening the healthcare systems. Moreover, it can guide future research by highlighting methodological trends, ultimately leading to improved work environments, quality of care, and patients’ oral health care outcomes.

## Acknowledgement

We would like to thank Sshaskank Kannan and Jiahao Deng, Research assistants, at McGill University, for their assistance in preparing this manuscript and their involvement throughout the overview.

## Author contributions

PKT has conceptualized the review. She has drafted the manuscript. All authors have substantially edited the manuscript. All authors have read and approved the manuscript.

## Data availability statement

They will be available upon request from the authors of the manuscript.

