## Appendix 2 for "Oral Healthcare Providers’ Well-being, Health and Experience (WHE) in Dental Care Settings: A Protocol for an Umbrella Review of Systematic Reviews and Meta-analyses"

### Ovid MEDLINE(R) ALL <1946 to November 08, 2024>

1 exp dental auxiliaries/ or exp dentists/ or practice patterns, dentists'/ or dentist's role/ 34881

2 (((dental or oral health* or oral medicine or oral maxillofacial) adj2 (assistant* or auxiliar* or clinician* or educator* or faculty or hygienist* or nurse* or paediatric* or pediatric* or personnel or practitioner* or profession* or provider* or researcher* or specialist* or staff* or student* or surgeon* or technician* or therapist* or work force or workforce)) or dental public health professional* or dentist or dentists or denturologist* or endodontist* or (oral adj3 surgeon*) or orthodontist* or periodontist* or prosthodontist* or dental education).ti,kf. 36369

3 (((dental or oral health* or oral medicine or oral maxillofacial) adj2 (assistant* or auxiliar* or clinician* or educator* or faculty or hygienist* or nurse* or paediatric* or pediatric* or personnel or practitioner* or profession* or provider* or researcher* or specialist* or staff* or student* or surgeon* or technician* or therapist* or work force or workforce)) or dental public health professional* or dentist or dentists or denturologist* or endodontist* or (oral adj3 surgeon*) or orthodontist* or periodontist* or prosthodontist* or dental education).ab,kf. /freq=2 30484

4 or/1-3 69890

5 exp dental auxiliaries/px or exp dentists/px 2910

6 adaptation, psychological/ or exp attitude of health personnel/ or dentist's role/ or exp stress, psychological/ 412176

7 quality of life/ or exp income/ or motivation/ or achievement/ or aspirations, psychological/ or conflict, psychological/ or goals/ or exp power, psychological/ 508025

8 mental health/ or psychological well-being/ 70324

9 health policy/ 74194

10 Work engagement/ or work-life balance/ or work performance/ or work schedule tolerance/ or workload/ 34332

11 (Attitude* or Perception* or Perspective* or Experience* or Reaction* or Adaptation* or Expectation* or Morale or Engagement or Role* or Accomplishment* or Achievement* or Satisfaction or Willingness or Motivation or Aspiration* or Goal* or Performance or Adoption or Behavio?r? or Implementation* or Compliance or Adherence or ((Mental or Psychological) adj health) or wellbeing or well-being or Wellness or Security or "Quality of life" or Burn-out or burnout or Fatigue or "Job security" or ((Financial or economic) adj status) or Income or Communicat* or Collaborati* or Partnership* or ((Patient* or famil* or caregiver*) adj health) or Empower* or (Health* adj2 (organi#ation* or system*)) or cultural safety or "Culture of safety" or Transparency or Discrimination or Support or "Diversity and Inclusion" or (Diversity adj3 Equity adj3 Inclusion) or Ethics or Morals or Values or Usability or Autonomy or environment or Implicit bias or Overt bias or Unconscious bias or Work-life balance or "Work schedule*" or Workload* or "work load*" or "Work* environment*" or "Work* condition*" or Professional relationship* or "Sense of meaning" or Compensation or HR policies or HR policy or Human resource* policies or Human resource* policy or (Health plan* adj1 implement*) or (Practice adj2 change*) or health policies or Health policy or public policies or Public policy or ((Health* system* or health care system* or management or organi#ation* or structural or administrati*) adj2 change*) or operation* or structur* or innovation* or restructur* or reform* or Responsibilit* or psychological stress* or stress level* or (work* adj3 stress*) or coping skill* or mental health or burden or turnover or work-related or workplace*).mp. or (bias or cultur* or job? or policies or policy or stress* or work).ti. 21591767

12 or/5-11 21620943

13 4 and 12 39305

14 limit 13 to "systematic review" 342

15 meta analysis.pt. or ((systematic adj2 review*) or (evidence adj (assessment* or review* or synthes*)) or meta analys* or meta?nalys* or (rapid adj2 (assessment* or review* or synthes*))).ti. 427712

16 13 and 15 425

17 14 or 16 492

https://proxy.library.mcgill.ca/login?url=https://ovidsp.ovid.com/ovidweb.cgi?T=JS&NEWS=N&PAGE=main&SHAREDSEARCHID=29ev69xGqZ0XmhFXLqd4QqGODj7K7sP56YGgYpBqBEb1pQKppDu8WkO20Inop97ZV
